# Comorbidities and Susceptibility to COVID-19: A Generalized Gene Set Meta-Analysis Approach

**DOI:** 10.1101/2020.09.14.20192609

**Authors:** MF Beckman, CK Igba, FB Mougeot, J-LC Mougeot

## Abstract

**Background:** The COVID-19 pandemic has led to over 820,000 deaths for almost 24 million confirmed cases worldwide, as of August 27^th^, 2020, per WHO report. Risk factors include pre-existing conditions such as cancer, cardiovascular disease, diabetes, obesity, and cancer. There are currently no effective treatments. Our objective was to complete a meta-analysis to identify comorbidity-associated single nucleotide polymorphisms (SNPs), potentially conferring increased susceptibility to SARS-CoV-2 infection using a computational approach.

**Results:** SNP datasets were downloaded from publicly available GWAS catalog for 141 of 258 candidate COVID-19 comorbidities. Gene-level SNP analysis was performed to identify significant pathways by using MAGMA program. SNP annotation program was used to analyze MAGMA-identified genes. COVID-19 comorbidities from six disease categories were found to have significant associated pathways, which were validated by Q-Q plots (*p*<0.05). The top 250 human mRNA gene expressions for SNP-affected pathways, extracted from publicly accessible gene expression profiles, were evaluated for significant pathways. Protein-protein interactions of identified differentially expressed genes, visualized with STRING program, were significant (*p*<0.05). Gene interaction networks were found to be relevant to SARS and influenza pathogenesis.

**Conclusion:** Pathways potentially affected by or affecting SARS-CoV-2 infection were identified in underlying medical conditions likely to confer susceptibility and/or severity to COVID-19. Our findings have implications in COVID-19 treatment development.

## INTRODUCTION

COVID-19 pandemic became prominent in Wuhan, China in December of 2019 [1]. As of August 27^th^, 2020, there have been over 24 million confirmed COVID-19 cases affecting over 200 countries [2]. This staggering number of cases includes more than 820,000 deaths, with the U.S. representing roughly one fourth of cases and deaths. WHO estimated the mortality rate of COVID-19 to be 3.4% in March 2020, which is significantly higher than reported average mortality rate of 0.1% for influenza [2].

COVID-19 is caused by severe acute respiratory syndrome coronavirus-2, SARS-CoV-2. This highly pathogenic coronavirus can cause severe respiratory illness and is highly contagious. The incubation period for SARS-CoV-2 can last up to 14 days with a median range of 4 to 5 days from exposure to onset of symptoms [3]. Transmission of the infection is due to inhalation of droplets and contact with contaminated surfaces. Symptoms include but are not limited to fever, cough, shortness of breath, fatigue, and body aches [4]. Current treatment for severe SARS-CoV-2 illness involves the management of complications associated with the disease and supportive care [1]. There is currently no treatment proven to be sufficiently effective for COVID-19 patients, although many drugs are being evaluated for effectiveness in reducing disease progression, severity, or mortality [5].

Multiple studies have shown that comorbidities influence severity, quality of life, or 1-year mortality, or altogether, in patients subject to viral infections [6, 7]. Risk factors for severity of SARS-CoV-2 infection include age 65 or above, and/or having a pre-existing condition [8, 9]. Thus, patients with pre-existing cardiovascular disease, diabetes, kidney dysfunction, obesity, and pulmonary diseases may have worse clinical outcomes when infected with SARS-CoV-2 [10].

Understanding pathways which could determine COVID-19 degree of susceptibility and severity is critical for drug development. Alternative drug combinations might act in synergy to complement future vaccination strategies, because no vaccination provides absolute protection [11]. Indeed, protection from vaccination against viral diseases may range from 9 to 90% and up to 60% of people vaccinated against influenza, for example, still fall ill due to the virus [12, 13]. In addition, vaccination is not systematically mandatory across countries.

Computational approaches may be utilized to identify candidate target molecules for the development of tailored drug treatments [14-16]. Insofar, drug targets can be predicted based on analysis of gene expression or genetic polymorphism profiles accessible through publicly available databases such as Gene Expression Omnibus (GEO) [17, 18]. Computational tools may (i) help elucidate or compare the mechanism(s) of action of drugs, (ii) assist in identification and characterization of interactions between a drug and its target, and (iii) provide a better understanding of mechanistic cellular reactions occurring at the molecular level in a drug response [19].

Computational approaches may also serve to investigate viral susceptibility and contribute to the development of vaccines [20-22]. For instance, machine learning has been used in the development of viral vaccines *(e.g*., influenza) and in the investigation of genetic adaptation of the virus to the host [23]. Indeed, the genetic variability of SARS-CoV-2 will likely impact vaccine development in future outbreaks. Additionally, an appropriate computational model that accounts for the complexities of molecular interactions in COVID-19 patients affected by comorbidities, would allow for a predictive and better assessment of viral mechanisms and patients’ response to drug or vaccine treatment.

The aim of this study was to complete a meta-analysis to identify genes associated with comorbidities/underlying medical conditions, potentially conferring increased susceptibility to SARS-CoV-2 infection or leading to the manifestation of more severe viral symptoms. To this end, we conducted a generalized gene set analysis using single nucleotide polymorphisms (SNPs) data from genome-wide association studies (GWAS) of a comprehensive list of possible comorbidities using Multi-marker Analysis of GenoMic Annotation (MAGMA)[24]. This analysis was complemented with (i) the investigation of predicted effects from the significant SNPs identified by MAGMA and (ii) the determination of differential human gene expression, most likely relevant to the pathogenesis of the viral respiratory illnesses, severe acute respiratory syndrome (SARS) and influenza.

## METHODS

### Multi-marker Analysis of GenoMic Annotation (MAGMA)

#### GWAS catalog and gene mapping

From an initial list of 258 mostly chronic diseases (data not shown), possibly representing comorbidities/underlying medical conditions associated with increased SARS-CoV-2 infectivity or disease severity, SNP datasets from the online GWAS catalog database [25] were identified using disease name, then downloaded and parsed. SNPs were mapped to genes using MAGMAv1.07b with the publicly available gene reference file NCBI37.3.gene.loc (https://ctg.cncr.nl/software/magma) [24].

#### Determination of multiple SNPs significance

The significance of SNPs (*p*-values) and derived sample sizes pertaining to genetic studies of comorbidities were extracted from the GWAS catalog datasets to compute correlations between neighboring genes and gene-level metrics *via* MAGMAv1.07b. To this end, the publicly available 1000 Genomes datasets (https://ctg.cncr.nl/software/magma) were used as reference files, considering the ethnicities associated with the possible comorbidity tested (European, East Asian, African, South American) [24]. To perform the gene set SNP analysis in MAGMAv1.07b, the ‘ncol’ flag was set to the to the sample size column in the SNP *p*-value file where each sample size corresponds to a p-value for the GWAS study completed. The flag for “multi=snp-wise” model was used to perform the ‘mean’ and ‘top1’ model analysis.

### Pathway analysis using Enrichment Map and MAGMAv1.07b programs

#### Reactome analysis

To conduct the pathway analysis, Reactome[26] human pathways were downloaded from Enrichment Map program [27] using Entrez gene IDs [28-30]. Computationally derived Gene Ontology (GO) biological terms and “No Data” were excluded. Based on the significant SNPs in comorbidity gene sets per MAGMAv1.07b analysis, significant Reactome gene ontology defined pathways were identified. The significant pathways also underwent a per-gene analysis with the MAGMAv1.07b model flag set to ‘alpha=0.05’ [24].

#### Interaction networks

Visualization of protein-protein interaction networks was completed using STRINGv11.0 [31] program by testing different confidence levels to identify ontologies of biological significance for the significant pathways associated with comorbidities.

#### Quality control

Possible comorbidity significant associated gene sets/pathways were checked for quality control by generating Quantile-Quantile (Q-Q) plots using observed quantiles and residual Z-scores of genes within the gene set, based on the MAGMAv1.07b publicly available Rv3.6.2 script (posthoc_qc_107a.r) [32, 33].

### Prediction of SNP effects

Ensembl’s Variant Effect Predictor program (VEP) [34] was used to analyze MAGMAv1.07b annotation files for each gene set associated with comorbidities [35]. MAGMAv1.07b annotation files were converted into VEP format using a bash script. All converted annotation files were uploaded into VEP online tool separately. VEP summary statistics and analysis tables were downloaded for the comorbidities’ associated genes and pathways found significant by MAGMAv1.07b. Corresponding tables were merged *via* Pythonv3.8.2 and SNPs containing a Sorting Intolerant from Tolerant (SIFT) score of 0 and a Polymorphism Phenotyping2 (PolyPhen2) score of 1, were removed (**Supplemental Data File 1**). Human Genome Organization (HUGO) gene symbols were extracted from the table with remaining SIFT and PolyPhen2 scores. Duplicate HUGO gene symbols were removed using Rv4.0.2. The most recently updated Affymetrix HG-U133A/B Human Genome Files [36, 37] containing annotated gene symbols and Entrez gene identifiers for all human genes were used to retrieve missing gene identification [38]. These tabular (.csv) files were merged and loaded into Rv4.0.2. Entrez gene IDs were matched to gene symbols from VEP analysis files to identify Affymetrix gene symbols. Genes and their corresponding Entrez ID’s were then matched to significant genes’ Entrez IDs found through combined MAGMAv1.07b - STRING analysis.

### Transcriptional gene expression analysis

GEO2R [39] was used to test the top 250 human mRNA gene expressions for each comorbidity based on available human data using NCBI GEO[39], by only including comorbidities that had significant pathways identified by MAGMAv1.07b and VEP STRING analyses. Human mRNA expression datasets comparing disease group to healthy controls since 2010 were searched. If no datasets were available post-2010 the latest dataset was downloaded using characteristics described for prior datasets. For diseases with no publicly available datasets comparing healthy controls to disease type, the newest, most relevant dataset was used. Tissue types used for analysis included: (i) peripheral blood mononuclear cells (PBMCs), (ii) cancer tissues, (iii) adipose tissue, (iv) pulmonary tissue, (v) post-mortem brain tissue, (vi) cardiovascular tissue, and (vii) blood stem cells.

For each comorbidity, human mRNA gene expression data corresponding to average log-fold change (aLFC) were formatted for clustering of genes identified by MAGMAv1.07b and VEP and subsequently matched to STRING protein-protein interactions. Gene weights were added manually to account for duplicate genes in the dataset. Genes were mean centered and normalized. Hierarchical clustering was completed using a similarity metric of Manhattan city-block distance for genes and arrays with average linkage *via* Cluster3.0v1.59. Clusters were visualized using heatmaps created using JavaTreeViewv1.1.6r4 [40]. Clustered groups of genes for MAGMAv1.07b and VEP genes were run separately through GeneCodisv4.0 online tool [41] for identification of possible biological processes or pathways involved in viral infection [42].

### Gene involvement in influenza and/or SARS

Significant genes (n=119) were investigated to determine their roles in relation to influenza and/or SARS respiratory viral infections. Genes were cross referenced using Pubmed [43] literature searches, DisGeNETv6 [44], Influenza Research Database[45] and conventional Google searches including HUGO gene symbol and either “influenza” or “SARS” [46, 47]. Risk of bias was assessed according to “Cochrane’s Handbook for Systematic Reviews of Interventions” [48]. Human tissue expression relevant to COVID-19 for genes with direct involvement was validated using Ensembl Expression Atlas [49, 50]. Genes not generally expressed in central nervous, cardiovascular, or pulmonary systems were removed from the dataset. Visualization of protein-protein interaction network of genes directly involved with influenza and SARS (caused by SARS-CoV-1) was completed using STRINGv11.0 using an interaction score of 0.400 [31].

## RESULTS

The overall computational analytical design and associated primary results are presented in **Figure 1**.

**Figure 1.**
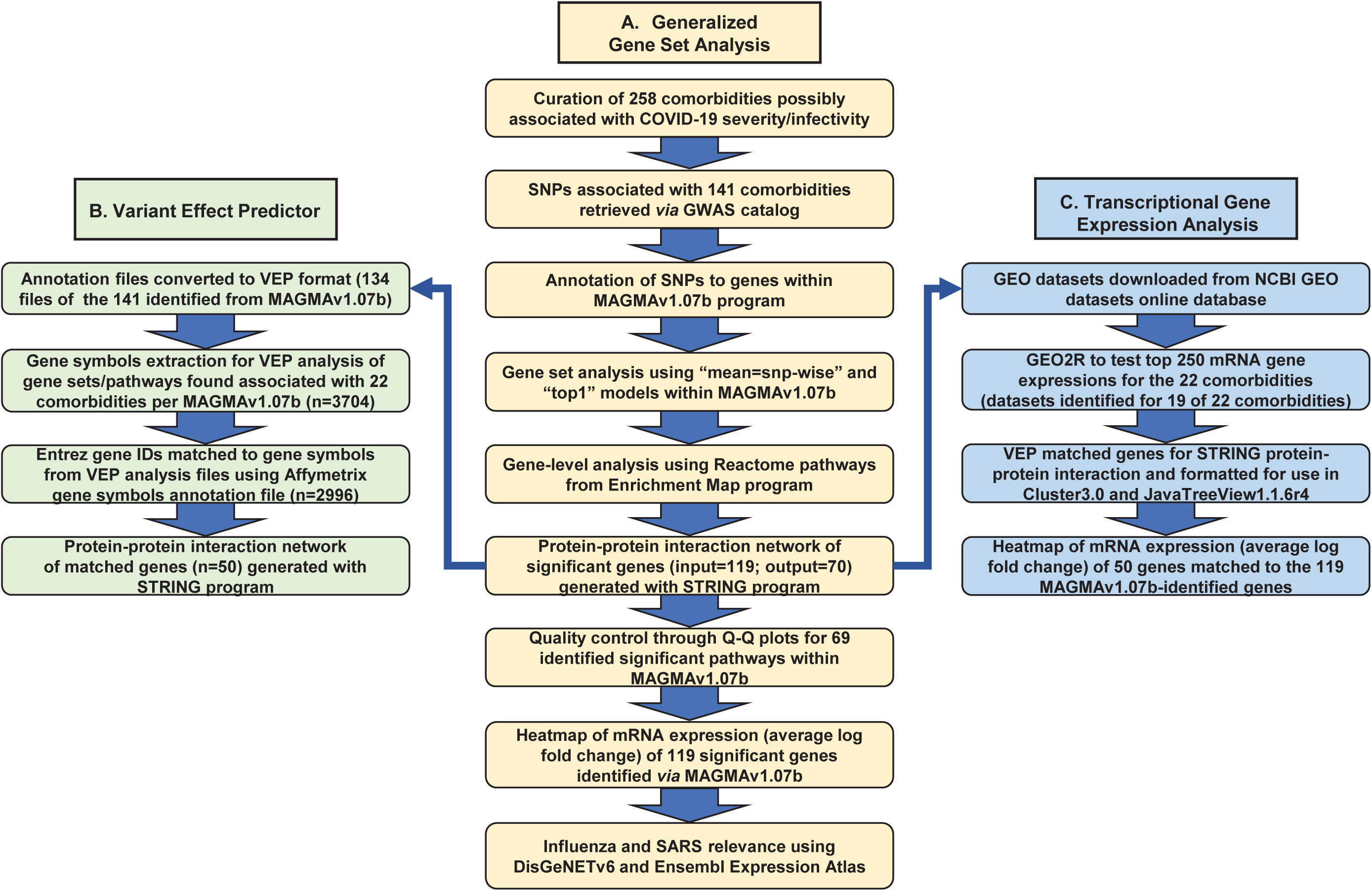
Computational analytical design for determination of genes/pathways associated with comorbidities, possibly contributing to COVID-19 severity/infectivity **Legend**. *Flowchart section A*. A list of candidate comorbidities (n=258) possibly associated with increased severity/infectivity of COVID-19 were curated. SNPs associated with comorbidities with available GWAS catalogdata (n=141) were analyzed. Multi-marker Analysis of GenoMic Annotation (MAGMA) was performed. SNPs were annotated to genes using NCBI gene reference file (NCBI37.3.gene.loc). In MAGMAv1.07b, gene set/pathway analysis was performed for which each SNP that was identified, using the “multi-mean=snp-wise” model-generated results, taking into account ethnicities associated with a possible comorbidity. Gene-level analysis was completed using Reactome pathways retrieved from Enrichment Map program. STRINGv11.0 protein-protein interaction program was used to visualize the network of 119 significant genes. Quantile-quantile (Q-Q) plots in Rv3.4.2 for 69 significant pathways were used for quality control. NCBI-gene expression omnibus (GEO) human mRNA differential expression datasets were downloaded *via* GEO2R for each comorbidity with associated genes/ pathways (n=19 of 22). Human mRNA expression was visualized with a heatmap of the 119 significant genes using Cluster3.0v1.59 and JavaTreeViewv1.1.6r4. Tissue expression relevance to SARS and influenza was determined using DisGeNETv6 and Ensembl Expression Atlas databases. *Flowchart section B*. MAGMAv1.07b annotation files were converted for Ensembl Variant Effect Predictor (VEP) format (n=134 of the 141 GWAS datasets). Gene symbols (n=3704) were extracted for VEP analysis from 22 significant comorbidity-associated genes/pathways per MAGMAv1.07b analysis. Entrez gene IDs (n=2996) were matched to gene symbols using Affymetrix gene symbols annotation files (HG-U133A/B Human Genome Files). STRINGv1.0 protein-protein interaction program was used to visualize the network (n=50 genes). *Flowchart section C*. NCBI-Gene expression omnibus (GEO) human mRNA differential expression datasets were downloaded *via* GEO2R for each of the 22 comorbidities with associated genes/pathways (n=19 of 22). VEP genes were matched to network genes and formatted for Cluster3.0v1.59. Human mRNA expression was visualized with a heatmap using average log-fold changes (aLFC) in JavaTreeViewv1.1.6r4 *(i.e*., 50 VEP identified genes matched to 119 MAGMA identified genes).

### MAGMA analysis of multiple SNPs associated with candidate COVID-19 comorbidities

To conduct generalized gene set analysis, we retrieved publicly available GWAS catalog datasets for 141 out of 258 COVID-19 possible comorbidities/underlying medical conditions. The 141 comorbidities were grouped into 8 categories by disease type based on organ most affected (**Table S1**). Following our MAGMA analysis (**Figure 1:** Flowchart section A), gene set and Reactome gene level analyses yielded 69 pathways representing 119 significant genes (p<0.05). These pathways were significant for 22 COVID-19 comorbidities representing 6 disease categories, namely, cancer (n=9); cardiovascular (n=4); neurologic/mental (n=3); respiratory (n=2); skin/musculoskeletal (n=1); autoimmune/endocrine/metabolic (n=3). Reactome significant pathways and genes obtained through MAGMAv1.07b gene-level analysis from Enrichment Map are shown in **Tables 1a and b**.

Using STRINGv11.0 program with the highest confidence interaction score (CIS) of 0.9, processing of the 119 genes yielded a protein-protein interaction network of 70 genes, which was found to be highly significant based on hypergeometric test with Benjamini-Hochberg correction (p=4.36×10^-11^) (**Figure 2a**). The top Kyoto Encyclopedia of Genes and Genomes (KEGG) pathway, identified by using STRINGv11.0, corresponded to Epstein-Barr virus infection with a false discovery rate of 6.72×10^-9^.

**Figure 2.**
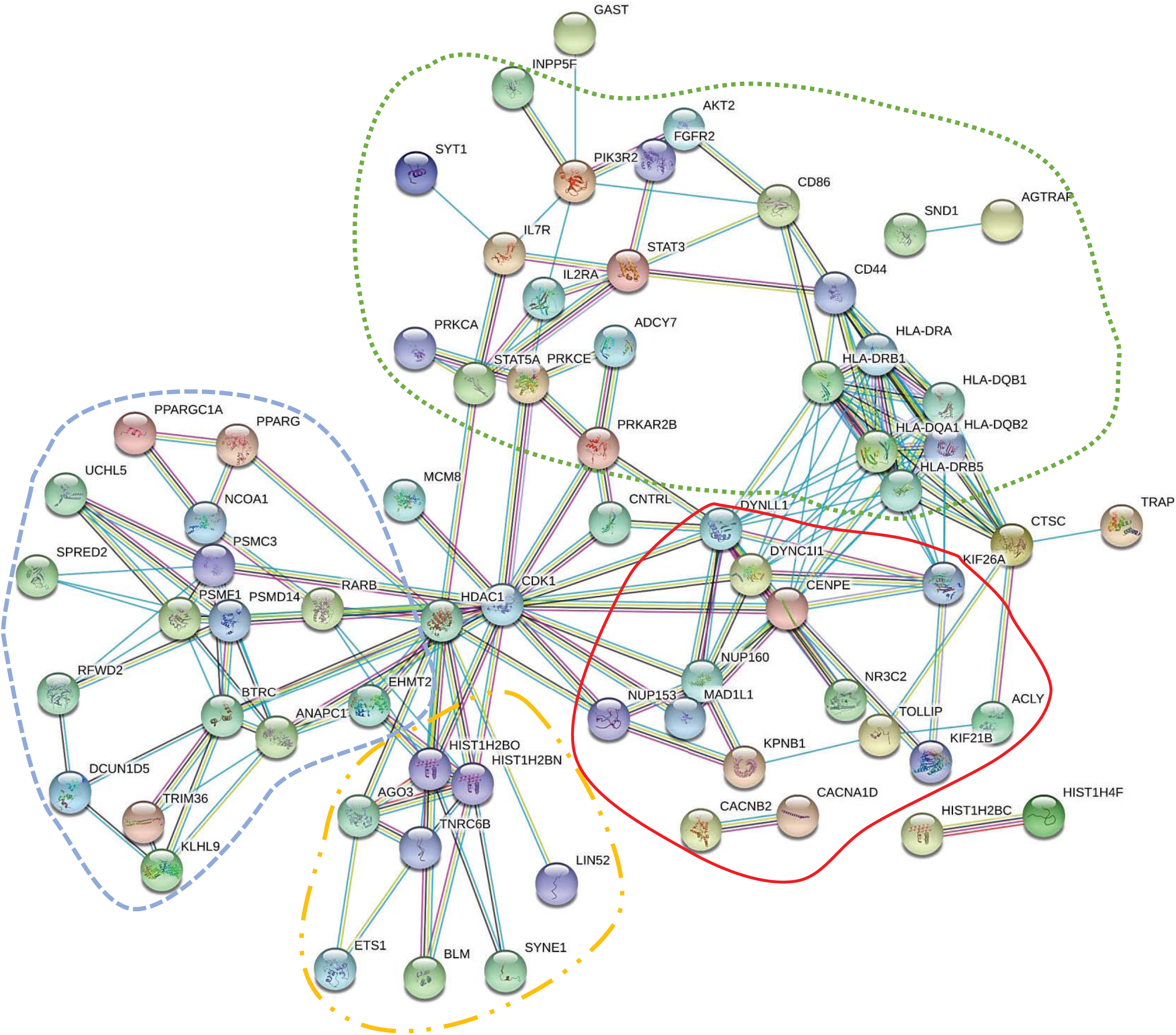
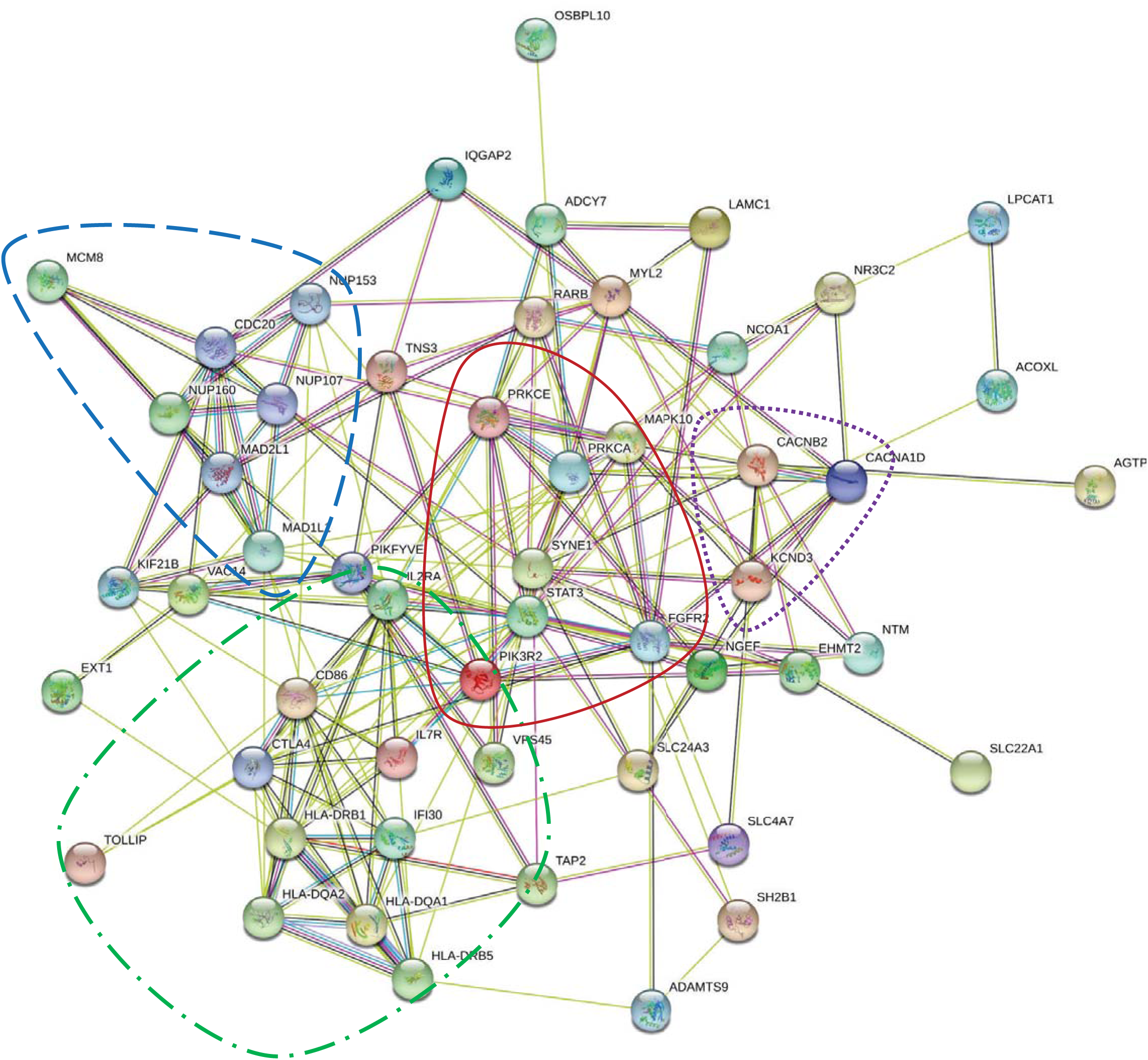
STRING protein-protein interaction network for significant COVID-19 comorbidity-associated genes *a. MAGMA significant genes (CIS=0.9)* *b. Matched MAGMA and VEP significant genes (CIS=0.15)* **Legend**. a. Significant genes were identified *via* MAGMAv1.07b gene-level analysis (https://ctg.cncr.nl/software/magma) using Reactome human pathways (https://reactome.org) obtained from Enrichment Map (http://baderlab.org/Software/EnrichmentMap) for 22 COVID-19 possible comorbidities with 119 significant genes. In STRINGv11.0 program, the confidence interaction score (CIS) was set to the maximum of 0.9, resulting in a network of 70 connected genes. Clustering by biological functions is represented by outlines. **Green (…):** cell regulation and immune response. **Red (––)**: cell transport and nervous tissue function. **Blue (--):** protein homeostasis and gene expression. **Orange (-..-):** transcriptional regulation and RNA-mediated silencing. b. Significant genes (n=50) were identified for 22 COVID-19 possible comorbidities in both VEP SNP and MAGMAv1.07b gene-level analyses. CIS was set to low value of 0.150, which resulted in integration of all 50 genes in a network of 55 genes. Clustering by biological functions is outlined. **Red (––):** antigen specific immune response. **Blue (--):** cell division and molecule formation/development. **Green (--.--):** cell growth, survival, proliferation, motility, and morphology. **Purple (…):** voltage gated ion channel transmembrane proteins. Note: Three gene symbols had other synonymous gene symbols (HLA-DRB1 and HLA-DRB5, HLA-DQA1 and HLA-DQA2, PIK3R2 and IFI30), which were also entered into STRINGv11.0 program for verification.

Verification of significant pathways using Q-Q plots showed a high association between genes and their relative gene ontology defined pathways, since all plots show a distribution of residual z-scores deviating from the diagonal early on. There were no Q-Q plots with any ambiguous feature. Significant genes had high levels of association with each pathway. Q-Q plots of more than five genes, representing the pathways ontologies “post-translational protein modification”, ”translocation of ZAP-70 to immunological synapse”, “metabolism” and “cell cycle” and associated possible COVID-19 comorbidities (including asthma), are described in **Figure S1**.

### VEP analysis of MAGMA-identified COVID-19 comorbidity-associated genes

Annotation files were converted for 134 of the 141 comorbidities with GWAS catalog datasets available (**Figure 1**: Flowchart section B). Of 3704 HUGO gene symbols extracted from VEP, 2996 corresponding Entrez gene IDs were identified using Affymetrix human genome annotation file. Of these gene IDs, 50 were matched with the 119 significant genes identified by MAGMAv1.07b for the 22 comorbidities with significant pathways (**Table 2**). Of the 50 genes, all were included in a protein-protein interaction network of 55 genes using a low CIS in STRINGv11.0 (**Figure 2b**). The top KEGG pathway identified using STRINGv11.0 was HTLV-1 infection with a false discovery rate of 4.38×10^-7^ using hypergeometric test with Benjamini-Hochberg correction.

### Transcriptional gene expression analysis of MAGMA- and VEP-identified genes

GEO human mRNA expression datasets were retrieved for 19 of 22 comorbidities. A description of GEO datasets is presented in **Table S2**. Using 119 MAGMAv1.07b identified genes (**Figure 1**: Flowchart section A), JavaTreeViewv1.1.6r4 clustered 4 of 9 cancer types in the heatmap (partial view in **Figure 3a**, full heatmap in **Supplementary Image**). Also, interstitial lung disease, multiple sclerosis, asthma, obesity, and heart failure were clustered (**Figure 3a**). VEP STRING matched genes (n=50) also clustered 4 of 9 cancer types and clustered interstitial lung disease, multiple sclerosis, asthma, obesity, and heart failure together (**Figure 3b**). In both heatmaps Nucleoporin 160 (NUP160), Nucleoporin 153 (NUP153), Fibroblast Growth Factor Receptor 2 (FGFR2), and Karyopherin Subunit Beta 1 (KPNB1) showed lower aLFC expression in 12, 10, 6, and 10 out of 19 comorbidities, respectively (**Figures 3a and b**). GeneCodisv4.0 was able to confirm the four above mentioned genes, as well as others to be involved in lung or viral biological processes.

**Figure 3.**
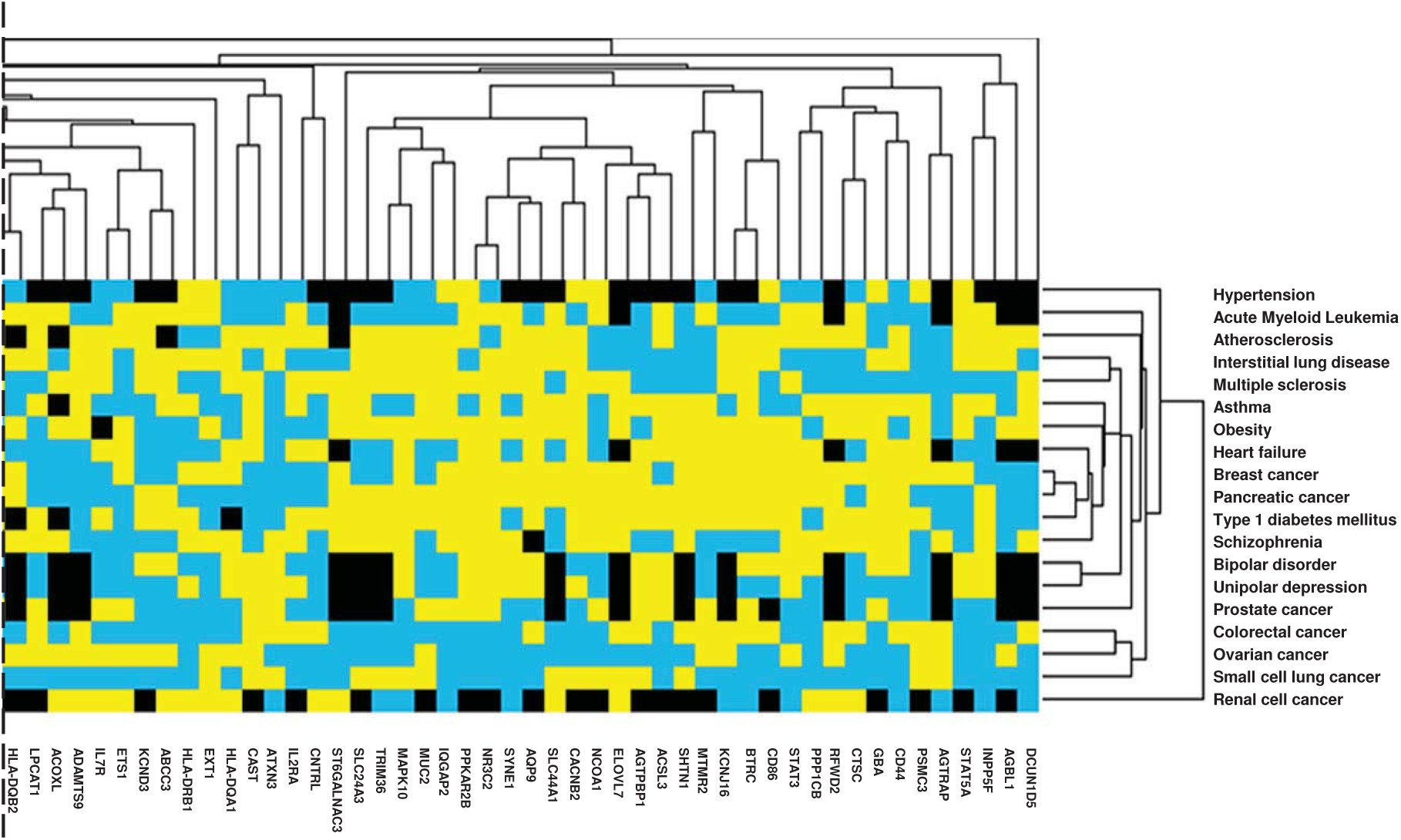
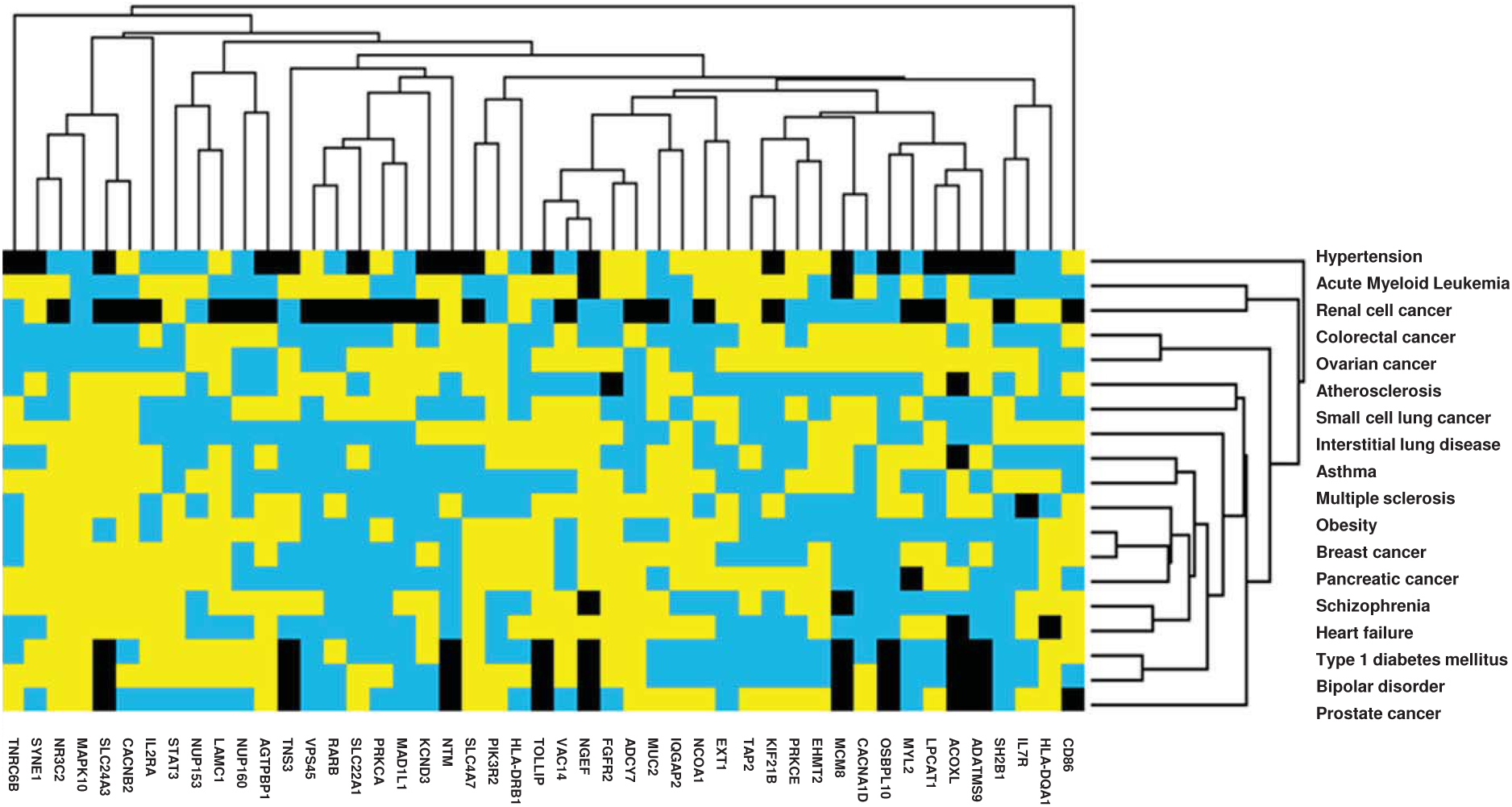
Human gene expression heatmap of MAGMA and VEP genes of COVID-19 associated comorbidities *a. Heatmap of MAGMAv1.07b significant genes (partial view)* *b. Heatmap of VEP matched genes* **Legend**. Heatmap from GEO2R human gene expression datasets using average log-fold changes (aLFC) from (**a**) 119 significant genes (p<0.05) identified *via* MAGMAv1.07b and (**b**) 50 genes matched to MAGMAv1.07b 120 significant genes from Ensembl Variant Effect Predictor (VEP) with 19 of 22 significant comorbidities identified via MAGMAv1.07b as arrays (**a**, **b**). Genes were filtered by removing those with less than 60% values present, mean centered and normalized. Hierarchical clustering was completed using weights for duplicate/synonymous genes with a similarity metric of city-block (Manhattan) distance for genes and arrays with average linkage through Cluster3.0v1.59 software. Heatmaps were created using JavaTreeViewv1.1.6r4. Yellow depicts positive aLFC, blue depicts negative aLFC, black depicts missing data and values of zero.

The 119 genes analyzed for gene expression were also investigated for their possible role in influenza and SARS-Cov-1 infection, as these might be relevant to SARS-Cov-2 infection. We identified three genes with a primary role in influenza infection: FGFR2, KPNB1 and NUP153 [51-54]. We also identified three genes KPNB1, Signal Transducer and Activator of Transcription 3 (STAT3), and Interleukin 2 Receptor Subunit Alpha (IL2RA) shown to play a significant role in SARS [55-57]. Genes identified as being possibly directly associated with influenza and/or SARS are shown in **Table S3**. STRING protein-protein interaction network yielded 38/46 (82.6%) genes involved in influenza and 15/17 (88.2%) genes involved in SARS, using an interaction score of 0.4 (**Figures 4a and b**). No GWAS study was found for SARS-CoV-1 infection to identify possible susceptibility genes within the 119 genes. Additionally, no studies were found to be at high risk for bias (**Table S4**).

**Figure 4.**
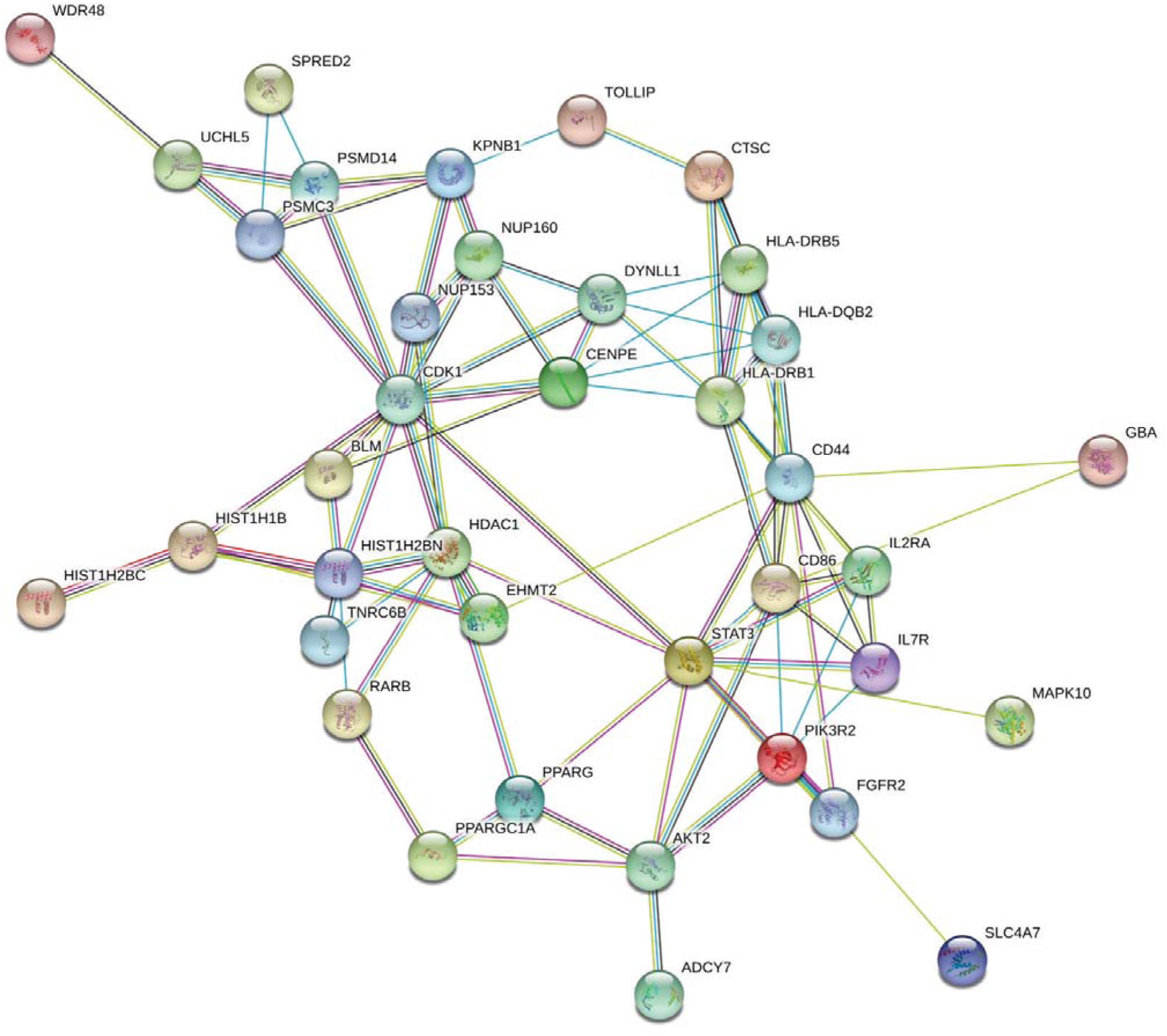
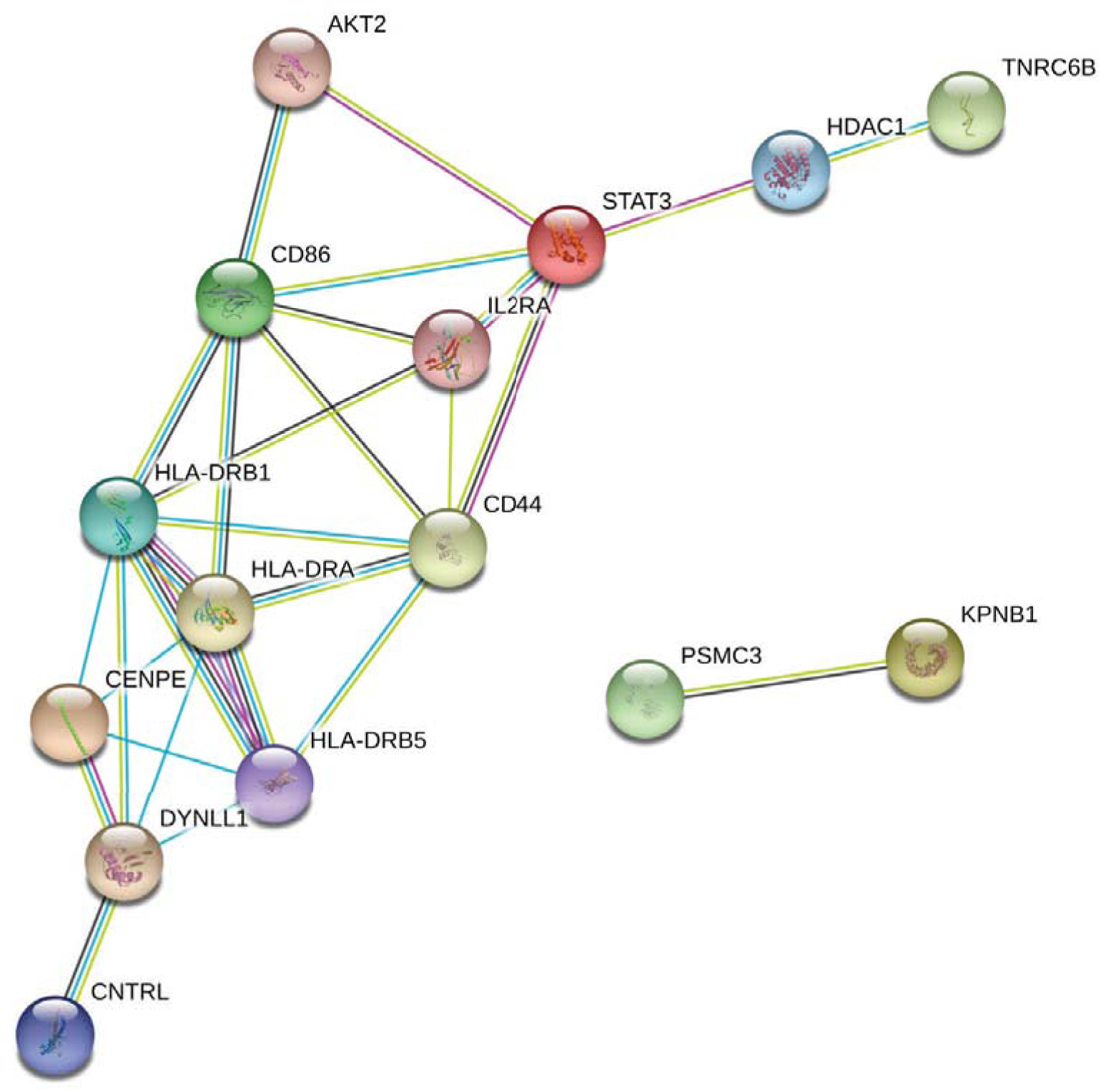
STRING protein-protein interaction of COVID-19 comorbidity-associated genes with involvement in influenza and/or SARS *a. Influenza (CIS=0.4)* *b. SARS (CIS=0.4)* **Legend**. STRING protein-protein interactions of MAGMAv1.07b identified genes with direct involvement with (**a**) influenza (n=46) and/ or (**b**) SARS (n=17) are shown. Level of stringency in STRINGv11.0 program was set to a medium confidence interaction score (CIS) of 0.4 in both influenza and SARS related molecular networks (**a**, **b**), resulting in a cluster of 38/46 (82.6%) and 15/17 (88.2%) genes, respectively.

## DISCUSSION

This is the first study conducting generalized gene set analysis on a broad spectrum of possible COVID-19 comorbidities, with the prospect of identifying comorbidity-specific genes that could impact infection by SARS-Cov-2.

Starting with a list of 258 diseases, our MAGMA pipeline was able to identify 69 significant Reactome pathways with a total of 119 significant genes corresponding to 22 comorbidities that might have implications in predicting the severity of SARS-CoV-2 infection (**Figure 1, Table 1, Table S1**). Of the 22 comorbidities, we were able to validate pathways associated with cardiovascular disease, diabetes, obesity, and pulmonary diseases. Cardiovascular diseases identified included heart failure, atherosclerosis, Kawasaki’s disease, and hypertension. Pulmonary diseases included asthma and interstitial lung disease. Cancer has been reported as a possible risk factor for COVID-19 [9]. We were able to identify nine cancers with GWAS data and significant associated pathways including acute myeloid leukemia, renal cell cancer, small cell lung cancer, and lung cancer. Furthermore, the known COVID-19 comorbidities, hypertension, obesity and diabetes had significant pathways and genes.

**Table 1.**
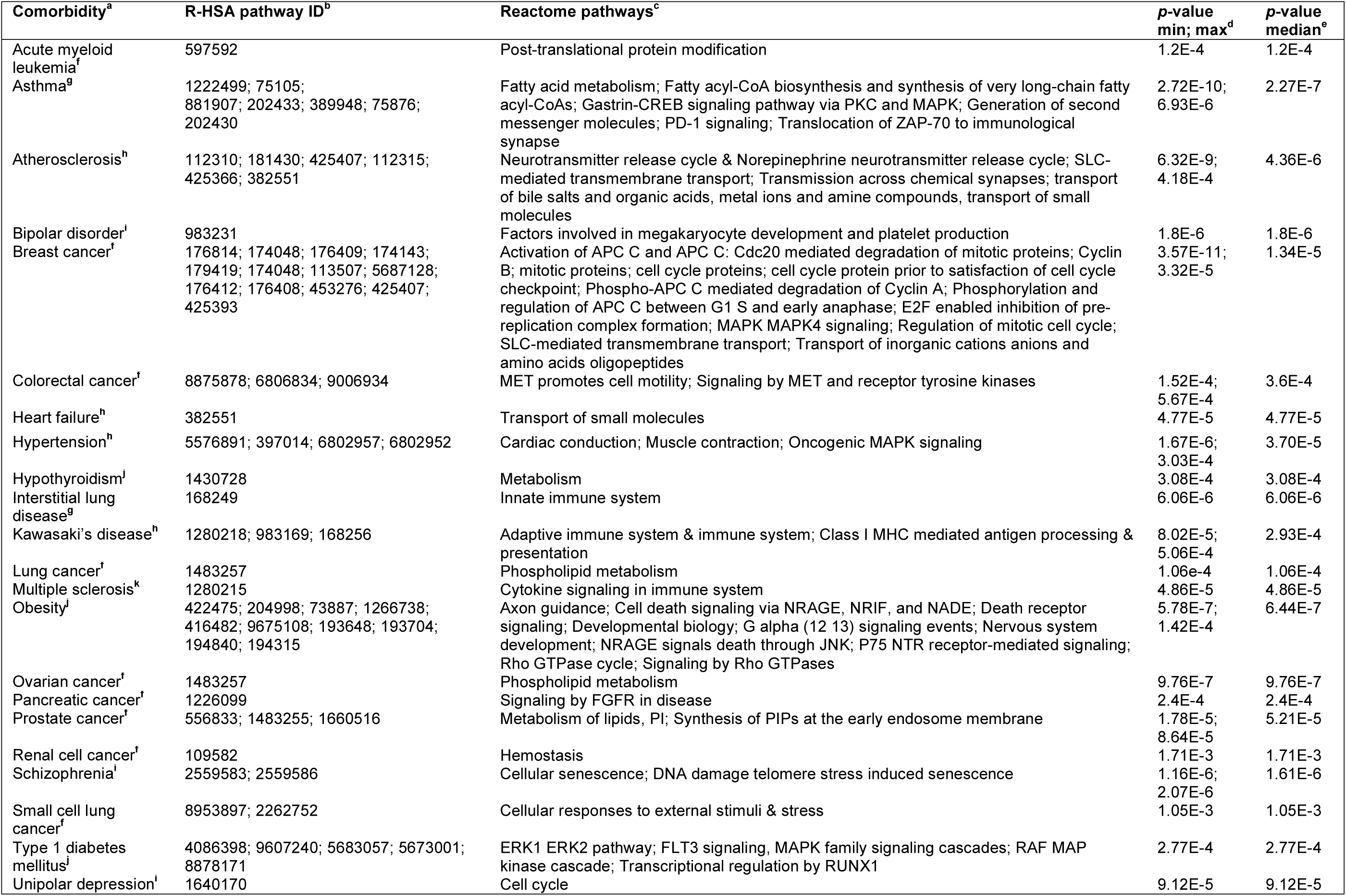
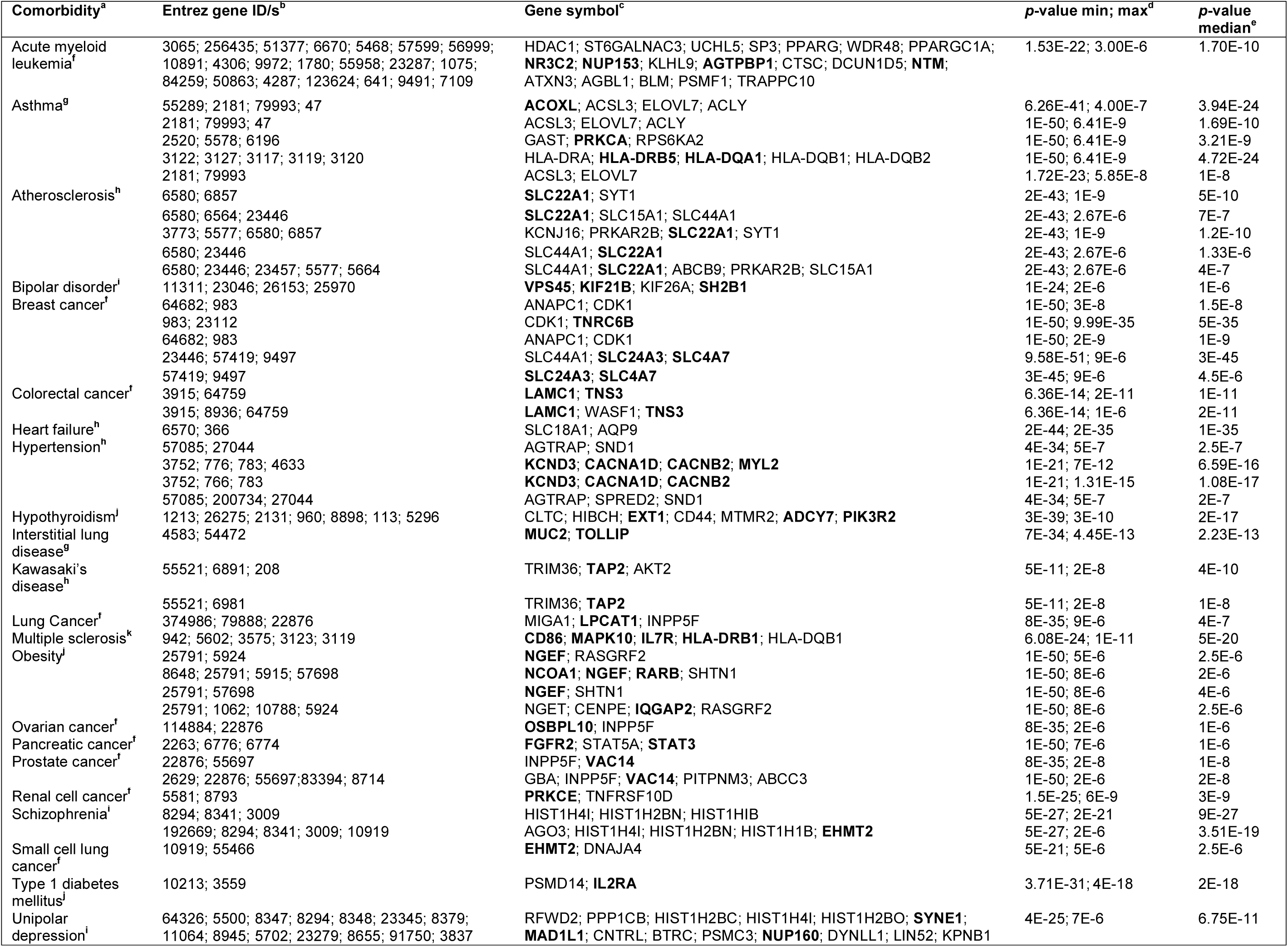
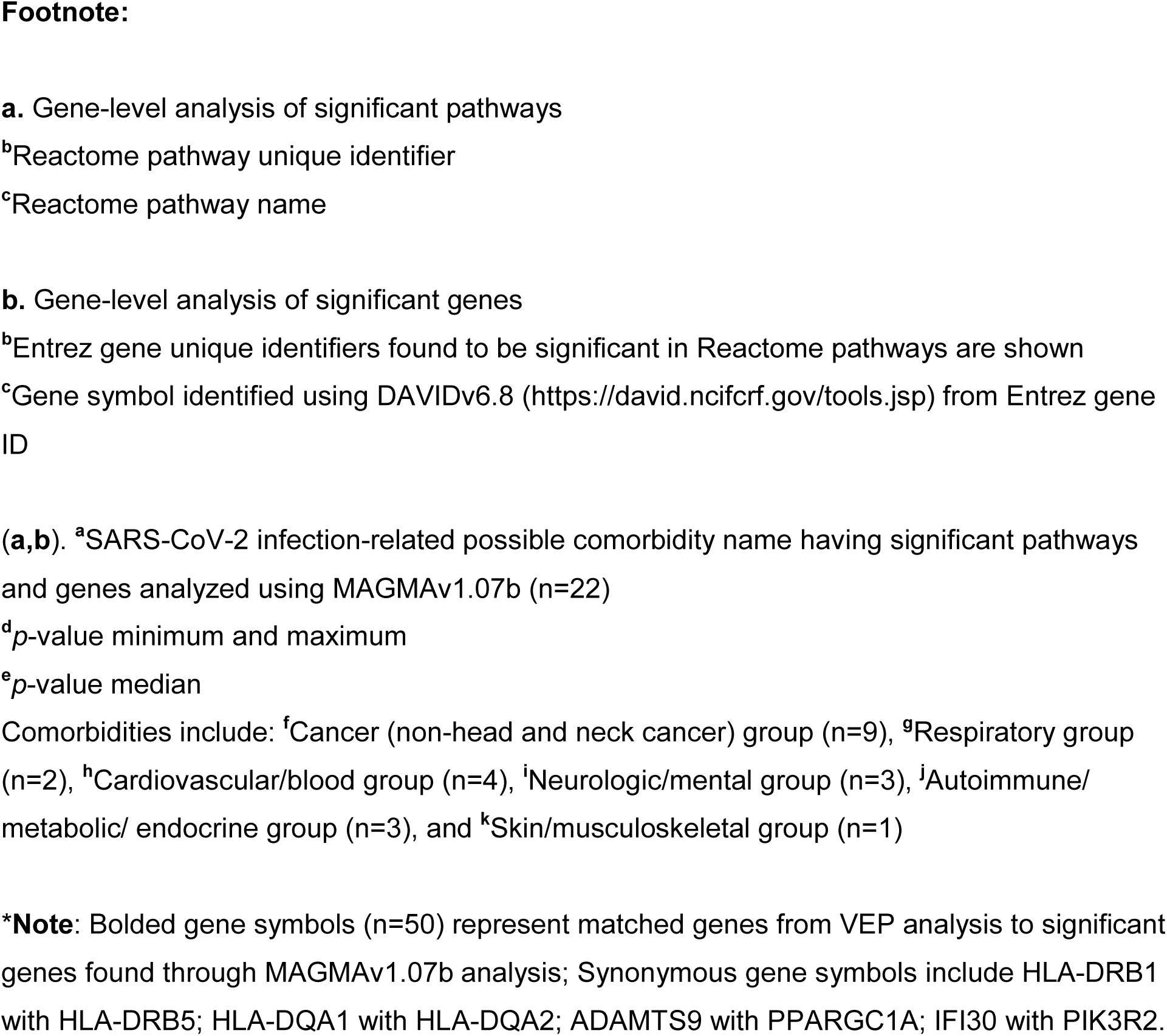
Reactome significant COVID-19 comorbidity-associated pathways and genes from EnrichmentMap program via MAGMAv1.07b. **a.** Gene-level analysis of significant pathways **b.** Gene-level analysis of significant genes

While Q-Q plots indicated validity of our findings, caution for interpretation of Q-Q plots must be used as these plots are normally used for pathways containing many genes. To a certain degree, these allow us to convey a certain level of confidence that there is a true association between gene and pathway [33]. In our analysis, however, less genes identified allowed us to narrow possible gene targets and pathways. Indeed, certain genes identified in our study may have significant biological relevance to infection by SARS-COV-2. For instance, sialyl transferase ST6 N-acetylgalactosaminide alpha-2,6-sialyltransferase 3 (ST6GALNAC3) was found significant in the post-translational protein modification pathway (**Figure S1**). Another sialyl transferase, ST6GALNAC1, has been previously investigated as a drug target against infection of smooth airways epithelial cells by influenza virus [58]. It remains, however, to be determined whether ST6GALNAC3, generally expressed at high levels in renal cell cancer[59], plays a significant role in COVID-19 pathogenesis. Interestingly, aLFC gene expression of ST6GALNAC3 was positive in our analysis for 8 of 19 comorbidities (including renal cell cancer), namely, in tumor, pulmonary, brain, adipose tissues and PBMCs (**Figure 3a&b**).

STRINGv11.0 analysis produced significant enrichment for both MAGMAv1.07b genes and VEP matched genes containing SNPs that had characteristics of deleterious effects (**Table 2**). Therefore, we believe the interactions among genes from significant pathways from MAGMA and matched VEP genes are likely not due to chance and that these genes are biologically connected. Furthermore, STRINGv11.0 analyses identified top KEGG pathways including, Epstein-Barr virus pathway (MAGMA genes), and HTLV-1 pathway (VEP matched genes). STRING was able to cluster 70 genes into four functional groups among the 119 MAGMA significant genes: cell regulation and immune response, cell transport and nervous tissue function, protein homeostasis and gene expression, transcriptional regulation and RNA-mediated silencing (**Figure 2a**). Additionally, NUP160, NUP153, and KPNB1 clustered tightly together in the cell transport and nervous tissue function group.

**Table 2.**
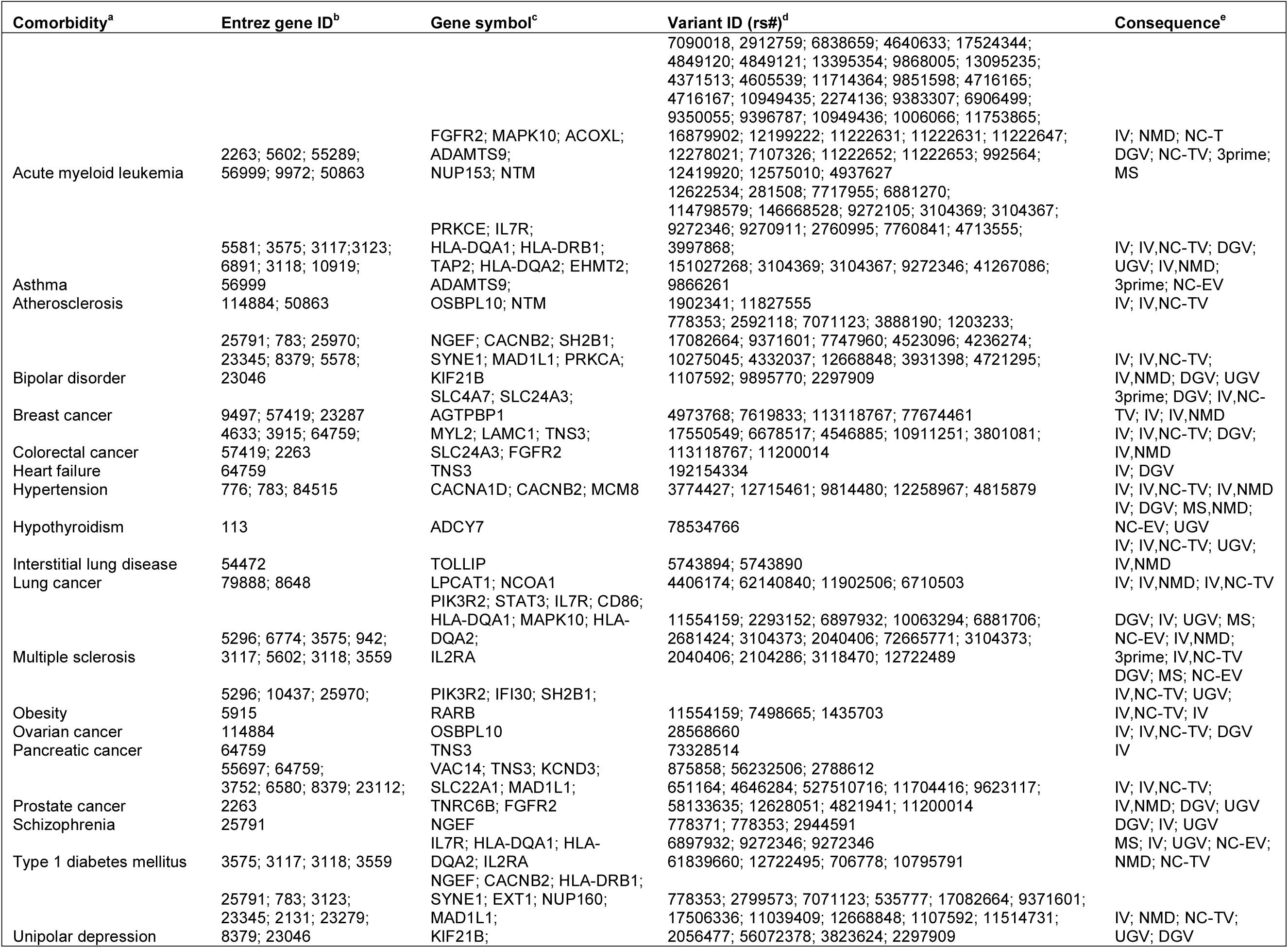
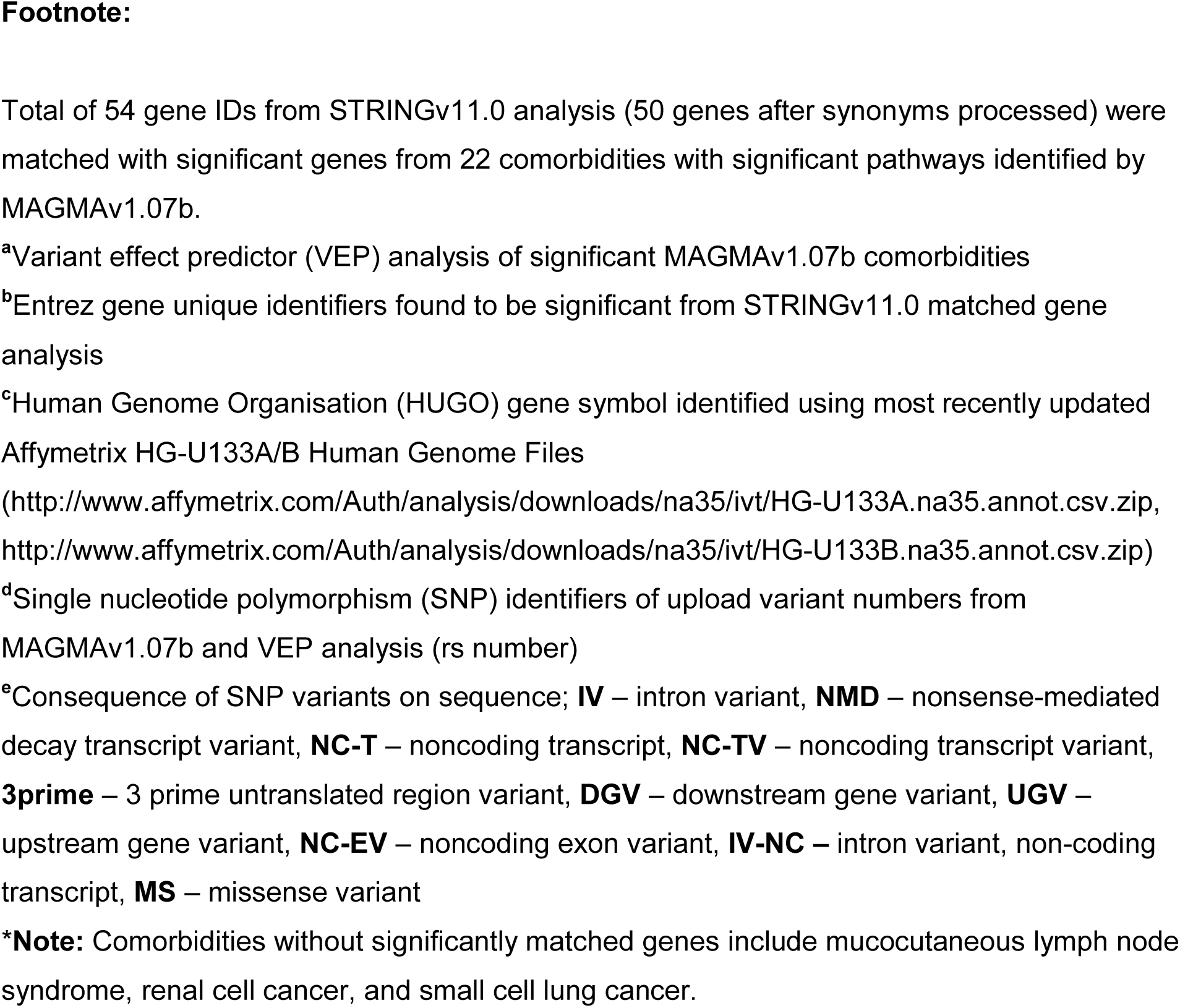
VEP genes and SNPs matched to significant MAGMAv1.07b COVID-19 comorbidity-associated genes.

STRINGv11.0 analysis of the 50 VEP matched genes with a lower confidence interval of 0.150 was required to obtain sufficient network connections for interpretation. Although network analysis may be subjective and is dependent on established knowledge, it is important to note that the enriched protein-protein interaction p-value was statistically significant. For the VEP matched gene STRINGv11.0 analysis, there were four distinct biological groupings recognized within the mapped network based on the closeness of protein interactions (**Figure 2b**). Those groupings were (i) antigen specific immune response, (ii) cell division and molecule formation/development, (iii) cell growth, survival, proliferation, motility, and morphology, (iv) and voltage gated ion channel transmembrane proteins. Notably, one of the comorbidities with significant associated pathways, breast cancer, contained SNPs affecting Solute Carrier Family 4 Member 7 (SLC4A7) and Solute Carrier Family 24 Member 3 (SLC24A3) genes. These genes are involved with sodium, calcium, and potassium ion transport and play a role in the malignant progression of breast cancer [60]. In addition, Euchromatic Histone Lysine Methyltransferase 2 (EHMT2) was mapped within close protein interactions. EHMT2 is involved with post-translational histone modification and epigenetic transcriptional repression. The orthologous gene (G9A) in drosophila is related to viral infection and susceptibility [61]. EHMT2 has been associated with the asthma comorbidity [62]. These results could indicate that EHMT2 deregulation possibly confers susceptibility to COVID-19.

The heatmap of MAGMA and VEP matched STRINGv11.0 genes shows that in 6 out of 19 comorbidities (**Figure 1: Flowchart section C**), FGFR2 had low aLFC (**Figure 3a and b**). Epithelial signaling by fibroblast growth factors is required for effective recovery from lung injuries resulting from influenza infection [51]. Our analysis coincides with previous findings linking induced inactivation of FGFR2 with increased mortality and influenza-induced lung injury [51]. KPNB1 (**Figure 2a**), NUP153, and NUP160 (**Figures 2a and 2b**) clustered in our heatmap analysis and were observed to have negative aLFC in majority (>50%) of possible comorbidities (**Figure 3a and b**). Downregulation of these factors under the condition of a comorbidity/underlying medical condition might impair cellular function in the presence of an active replicating virus.

Indeed, it has been previously established that a lower concentration of KPNB1 (interacting with NUP153, **Figure 2a**) in epithelial colorectal adenocarcinoma cells results in diminished Signal Transducer and Activator of Transcription 1 (STAT1) nuclear translocation activity [55]. Such regulation is critical for STAT1-activated genes to determine the level of anti-viral response and disease severity in an infection by SARS-CoV-1 [55]. Furthermore, KPNB1 is involved in the early stage of influenza virus replication *via* nuclear trafficking, by way of, nuclear import of viral cDNA or viral/host proteins into the host chromosome [52, 53].

Based on previous studies, the interaction between NUP153 and KPNB1 has been investigated in relation to nuclear transport [63]. The degradation of NUP153 in influenza virus A infected cells, such as Madin-Darby canine kidney II and human lung epithelial cells, results in an enlargement and widening of nuclear pores [54]. This disease process allows viral ribonucleoprotein complexes to be exported from the nucleus to the plasma membrane[54]. Additionally, NUP160 has been shown to work in conjunction with NUP153 to mediate nuclear import and export [64]. Therefore, degradation of one or both can prevent the import of signal transducers and activators of transcription, reducing effectiveness of the anti-viral interferon response [65]. Our results support the interactions between these genes and viral respiratory diseases, such as influenza and SARS.

NUP153 showed negative aLFC of three cancer types (tumor tissues), three cardiovascular diseases (PBMCs and cardiovascular tissue), two respiratory diseases (pulmonary tissues), and two autoimmune/endocrine/metabolic diseases (PBMCs and adipose tissue). NUP160 showed lower aLFC in four cancer types (tumor tissues), two cardiovascular disease (PBMCs and cardiovascular tissues), two respiratory diseases (pulmonary tissues), two autoimmune /endocrine/metabolic diseases (PBMCs and adipose tissue), one mental health disease (post-mortem brain tissue), and one skin/musculoskeletal (PBMCs). KPNB1 showed low aLFC in four cancer types (tumor tissues), two respiratory diseases (pulmonary tissues), two mental health diseases (post-mortem brain tissue), one cardiovascular disease (PBMCs), and one autoimmune/endocrine/metabolic disease (adipose tissue).

Moreover, FGFR2 showed lower aLFC in four cancer types (tumor tissue), one respiratory disease (pulmonary tissues), and one cardiovascular disease (PBMCs) (**Figure 3a and b**). Notably, the possible comorbidities, asthma and prostate cancer, were identified as having all four genes *(i.e*., FGFR2, KPNB1, NUP153, and NUP160) downregulated. Three of the four genes were downregulated in obesity (KPNB1, NUP153, NUP160), heart failure (FGFR2, KPNB1, NUP153), and interstitial lung disease (KPNB1, NUP153, NUP160) (**Figure 3a**). Only NUP153, and NUP160 were downregulated in hypertension and type 1 diabetes mellitus (**Figure 3a and b**).

GeneCodis was able to identify FGFR2 as being involved in mesenchymal cell differentiation involved in lung development while NUP153 and NUP160 are involved in viral replication process and intracellular transport of viruses. STAT3 was identified as being involved in primary miRNA binding and viral process and has been observed to be downregulated in SARS-CoV-1 infected Vero E6 kidney epithelial cells extracted from an African green monkey [56]. Additionally, IL2RA has been recently identified as significantly upregulated in the plasma of patients with severe COVID-19 [57] (**Figures 2a and 2b**). In our analysis, GeneCodis also identified Transporter 2, ATP binding Cassette Subfamily B Member (TAP2), Major Histocompatibility Complex, Class II, DR Beta 1 (HLA-DRB1), and Major Histocompatibility Complex, Class II, DQ Beta 1 (HLA-DQB1) as being involved in Epstein-Barr virus infection. Further research is needed to confirm these genes (or associated regulations) as possible drug targets for SARS-CoV-2 infection.

### Limitations

While there is no shortage of publicly available data, not all diseases have the same level of dedicated research. Therefore, not all possible comorbidities had publicly available SNP datasets from GWAS catalog or human mRNA gene expression datasets from NCBI’s GEO datasets database. This resulted in a large decrease from 258 possible comorbidities to 141. Additionally, we were only able to use 19 of 22 significant comorbidities for GEO2R analysis and heatmap visualization. Another caveat is that GEO2R mRNA expression datasets have been generated through different independent studies using different genomic platforms and analysis pipelines, so that optimal normalization of raw data cannot be implemented. Little is still known about COVID-19 pathogenesis, although research on the matter has increased greatly since the beginning of the pandemic.

### Conclusions

Significant pathways were identified associated with comorbidities/underlying medical conditions conferring susceptibility and/or severity to SARS-CoV-2 infection, which have been reported in conjunction with decreased clinical outcomes. Our findings may have implications in development of COVID-19 therapies.

## Data Availability

The data for this study is available upon request.

## SUPPLEMENTAL MATERIALS

Supplemental File 1: Figure S1 – COVID Q-Q plots

Supplemental File 2: Table S1 – COVID Comorbidities

Supplemental File 3: Table S2 – GEO dataset description

Supplemental File 4: Table S3 – Influenza/SARS involved genes

Supplemental File 5: Table S4 – Risk of bias assessment

Supplemental File 6: Supplementary Image – Full Heatmap Figure 3a

Supplemental File 7: Supplementary Data File 1.csv

## ABBREVIATIONS

ADAMTS9: a disintegrin-like and metalloprotease with thrombospondin type 1 motif, 9
AHABE: adenocarcinoma human alveolar basal epithelial
aLFC: average log-fold change
A-MuLV: Abelson murine leukemia virus
C127: mouse fibroblast
CIS: confidence interaction score
COVID-19: coronavirus disease of 2019
CTL: cytotoxic T-lymphocyte
EHMT2: euchromatic histone lysine methyltransferase 2
FGFR2: fibroblast growth factor receptor 2
GEO: Gene Expression Omnibus
GIDEON: Global Infectious Disease and Epidemiology Network database
GO: Gene Ontology
GWAS: genome-wide association studies
HBE: human bronchial epithelial
HEK: human embryonic kidney
HLA-DRB1: major histocompatibility complex, class II, DR Beta 1
HLA-DRB5: major histocompatibility complex, class II, DR Beta 5
HLA-DQA2: major histocompatibility complex, class II, DQ Beta 1
HLA-DQB1: major histocompatibility complex, class II, DQ Beta 1
HTL: helper T-lymphocyte
HUGO: Human Genome Organization
IFI30: IFI30 lysosomal thiol reductase
IL2RA: interleukin 2 receptor subunit alpha
KEGG: Kyoto Encyclopedia of Genes and Genomes
KPNB1: karyopherin subunit beta 1
MAGMA: Multi-marker Analysis of GenoMic Annotation
MDCK: madin-darby canine kidney
miRDB: microRNA Target Prediction Database
ND: No Data
NUP153: nucleoporin 153
NUP160: nucleoporin 160
PBMCs: peripheral blood mononuclear cells
PIK3R2: phosphoinositide-3-kinase regulatory subunit 2
PM: primary macrophages
PPARGC1A: peroxisome proliferator-activated receptor gamma coactivator 1 alpha
PolyPhen2: Polymorphism Phenotyping 2
Q-Q: quantile-quantile
SARS: severe acute respiratory syndrome
SARS-CoV-1: severe acute respiratory syndrome coronavirus-1
SARS-CoV-2: severe acute respiratory syndrome coronavirus-2
SIFT: Sorting Intolerant from Tolerant
SLC4A7: solute carrier family 4 member 7
SLC24A3: solute carrier family 24 member 3
SNPs: single nucleotide polymorphisms
ST6GALNAC1: ST6-acetylgalactosaminide alpha-2,6-sialytransferase 1
ST6GALNAC3: ST6 N-acetylgalactosaminide alpha-2,6-sialyltransferase 3
STAT1: signal transducer and activator transcription 1
STAT3: signal transducer and activator of transcription 3
TAP2: transporter 2, ATP binding cassette subfamily B member
VEP: Variant Effect Predictor
Vero: epithelial kidney cells derived from African green monkey *(Chlorocebus spp.);*
WHO: World Health Organization
ZAP-70: zeta-chain associated protein kinase 70

## AUTHORS’ CONTRIBUTIONS

Dr. Jean-Luc Mougeot and Dr. Farah Bahrani Mougeot conceived the study, contributed to the design of the analytical strategy, data interpretation and verification. Micaela Beckman designed the overall analytical strategy, conducted most computational analyses and data interpretation, and wrote the manuscript draft. Chica Igba contributed to the design of analytical strategy, conducted analyses and participated into data interpretation. All authors had significant contributions in writing and revisions of main manuscript, tables and figures.

## ACKNOWLEDGEMENTS

We thank Kathleen Sullivan for her editorial expertise in preparation of this manuscript. This work was supported by the Atrium Health Foundation Research Fund (internal).

## AVAILABILITY OF DATA

Publicly available data were used. Analyzed data available upon request.

## CONSENT FOR PUBLICATION

All authors have approved the final version of this manuscript.

## ETHICS APPROVAL

N/A

